# Reaching collective immunity for COVID-19: an estimate with a heterogeneous model based on the data for Italy

**DOI:** 10.1101/2020.05.24.20112045

**Authors:** Andrey Gerasimov, Georgy Lebedev, Mikhail Lebedev, Irina Semenycheva

## Abstract

**Background:** At the current stage of COVID-19 pandemic, forecasts become particularly important regarding the possibility that the total incidence could reach the level where the disease stops spreading because a considerable portion of the population has become immune and collective immunity could be reached. Such forecasts are valuable because the currently undertaken restrictive measures prevent mass morbidity but do not result in the development of a robust collective immunity. Thus, in the absence of efficient vaccines and medical treatments, lifting restrictive measures carries the risk that a second wave of the epidemic could occur.

**Methods:** We developed a heterogeneous model of COVID-19 dynamics. The model accounted for the differences in the infection risk across subpopulations, particularly the age-depended susceptibility to the disease. Based on this model, an equation for the minimal number of infections was calculated as a condition for the epidemic to start declining. The basic reproductive number of 2.5 was used for the disease spread without restrictions. The model was applied to COVID-19 data from Italy.

**Findings:** We found that the heterogeneous model of epidemic dynamics yielded a lower proportion, compared to a homogeneous model, for the minimal incidence needed for the epidemic to stop. When applied to the data for Italy, the model yielded a more optimistic assessment of the minimum total incidence needed to reach collective immunity: 43% versus 60% estimated with a homogeneous model.

**Interpretation:** Because of the high heterogeneity of COVID-19 infection risk across the different age groups, with a higher susceptibility for the elderly, homogeneous models overestimate the level of collective immunity needed for the disease to stop spreading. This inaccuracy can be corrected by the homogeneous model introduced here. To improve the estimate even further additional factors should be considered that contribute to heterogeneity, including social and professional activity, gender and individual resistance to the pathogen.

**Funding:** This work was supported by a grant from the Ministry of Education and Science of the Russian Federation, a unique project identifier RFMEFI60819X0278.

## Introduction

Most countries in the world are currently severely affected by the COVID-19 epidemic ^1^ The undertaken anti-epidemic restrictions have been effective, but there remains the risk of the epidemic second wave after the restrictions are lifted and people return to their normal way of life. Indeed, the potential vaccines are only being explored ^2-4^, so mass vaccination of the population will not start any time soon ^5^. In these conditions, epidemic dynamics depend on the basic reproductive rate (that can be lowered by anti-epidemic measures) and the number of susceptible-exposed-infectious-recovered cases ^6,7^.

As the number of infections increases (slowly with restrictive measures), a population immunity state can be reached where repeated occurrences of infection only lead to dilapidated chains of successive infections, not mass infections ^8-10^. It is important to be able to forecast such an event when deciding to lift the anti-epidemic restrictions ^11,12^.

Here we developed a mathematical model for assessing the minimum incidence of COVID-19 needed to reach collective immunity, which would assure that the epidemic cannot restart the cessation of quarantine measures. The key feature of our model is that it is heterogeneous, that is it accounts for the differences in the infection risk across affected subpopulations. This is important for COVID-19 because the severity of this disease strongly depends on age ^1,13,14^ As such, our model offers more precise estimates compared to the commonly used homogeneous models.

## Research in context

### Evidence before this study

We conducted a search of MedRxiv, PubMed, and BioRxiv for articles published in English from inception to May 9, 2020, with the keywords “COVID-19”, “SARS-CoV-2”,“reproduction number”, “R0”, “age”, “homogeneous model”, and “heterogeneous model”. While this search yielded several useful references regarding COVID-19 modeling, the basic reproduction number of this disease, and age-related heterogeneity, we did not find an approach similar to ours to modeling COVID-19 dynamics and estimating the total incidence and population immunity. Therefore, our study provides a novel model that estimates these metrics accurately and with a minimal number of model parameters.

### Added value of this study

Our report the results obtained with a heterogeneous model, which is different from the commonly used homogeneous models of the epidemic dynamics. This is especially important for COVID-19, whose risks are strikingly age-dependent. With these modelling results, we contribute to the demanding issue of lifting anti-epidemic restrictions, the measure that could result in the second wave of the epidemic because of an insufficient level of collective immunity. In addition to assessing the minimum collective immunity level, our report provides a framework for incorporating additional sources of heterogeneity and for monitoring the epidemic dynamics for different subpopulations. As such, our model is an important tool that could be useful for making crucial decisions, which eventually will result in saving human lives.

### Implications of all the available evidence

Criteria for lifting restrictions (when and how) are critical for the overall success of anti-epidemic measures. Our contribution to this issue is the assessment of the level of collective immunity that assures that the disease would nor restart when the restrictions are lifted. The assessment was made using a heterogeneous model that accounted for the age-related differences in COVID-19 infection risk. Future research should consider additional sources of heterogeneity, such as heterogeneity associated with social and professional activity, gender and individual resistance to the pathogen.

## Methods

We reasoned our model the following way. The simplest Kermack-McKendrick susceptible–infected–recovered (SIR) model of an epidemic ^15^ assumes that each member of the population can be in one of three states: susceptible, infected, recovered (or immune), and the probability of transition between the states is the same for all members of the population. The epidemic dynamics is then described by the differential equations:

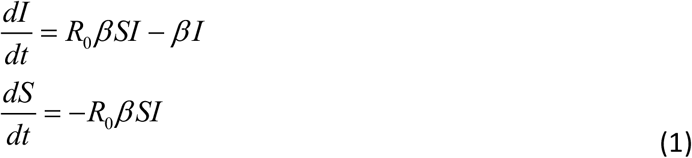

where *S* and *I* are susceptible and infected subpopulations, respectively. The term *R_0_* is the basic reproductive number, that is the average number of people infected by one infected person with 100% susceptibility. Furthermore, *βI* is the recovery intensity for the infected people, 1/*β* is the average duration of the disease, and *R*_0_*βSI* is the intensity of the infection flow.

This representation can be further improved by the susceptible-exposed-infectious-recovered (SEIR) model that accounts for the fact that there is a period when after being infected a person does not infect the others ^6,7^ Additionally, not only the contagiousness but also the probability of recovery (or death) depends on the time elapsed since the infection. To introduce these factors, let *J (t)* be the intensity of the emergence of new cases, *J = R*_0_*βSI*. Then the system dynamics are described by the equations:

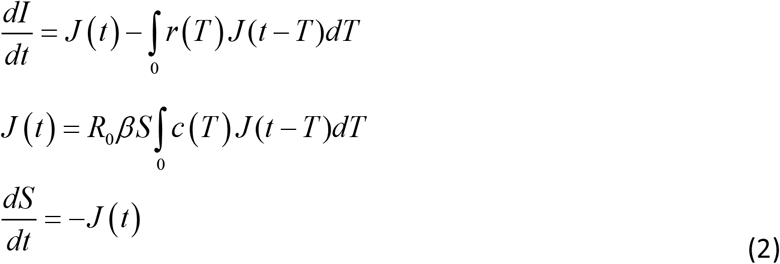

where *r (T)* is the probabilistic distribution of the disease duration before recovery or death 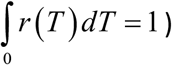, and *c (T)* is the dynamics of the dependence of the average contagiousness over the time elapsed since being infected 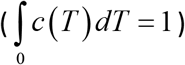.

Anti-epidemic activities like social isolation can be factored in by considering *R_0_* to be time-dependent rather than constant. Then, morbidity decreases if the recovery flow is higher than the infection flow: 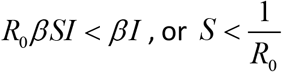.

Taking *R*_0_ = 2.5 as an estimate for the basic reproductive number, the susceptible subpopulation, *S*, should be less than 40% for the epidemic to decline. In other words, 60% of the population should contract the disease.

This assessment, however, overestimates the expected morbidity because it does not take into account the presence of low and high-risk groups. Let h be an individual risk of infection, so
that the average value of h for the population is 1 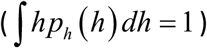, and *S (h, t), I (h, t)* and *J (h, t)* are the values for risk *h* at time *t*. Then the dynamics of the epidemic are described by the equations

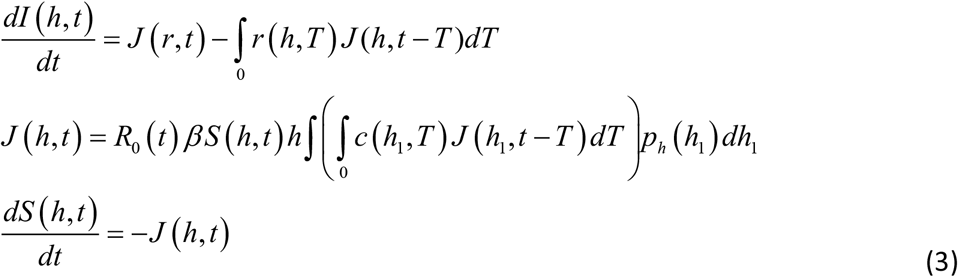

Since 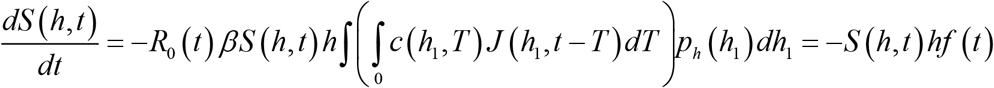 at the epidemic beginning, *S* (*h*,0) = 1, then

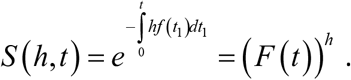

The condition for the epidemic decline is then: 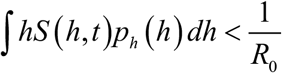

Here we propose using this heterogeneous-model approach for the assessment of COVID-19 population immunity level. Italy is taken as an example.

## Results

Since the risk of COVID-19 is highly dependent on age ^13,14^, we considered an epidemic model where the individual risk of infection was a function of age. At the initial stage of an epidemic the proportion of infected persons is low. Therefore, individual risk, *h*, is proportional to the number of infections in each age group, and the distribution of *h* can be estimated from the distribution of morbidity by age.

Table 1 presents this assessment for the data from Italy. We calculated *h* as being proportional to morbidity; the average *h* for the entire population is 1. It can be seen from the last column of the table that people over 80 have the risk that is 2.5 times higher than the average, while the risk in children and adolescents is 15 times lower than the average.

**Table 1.**
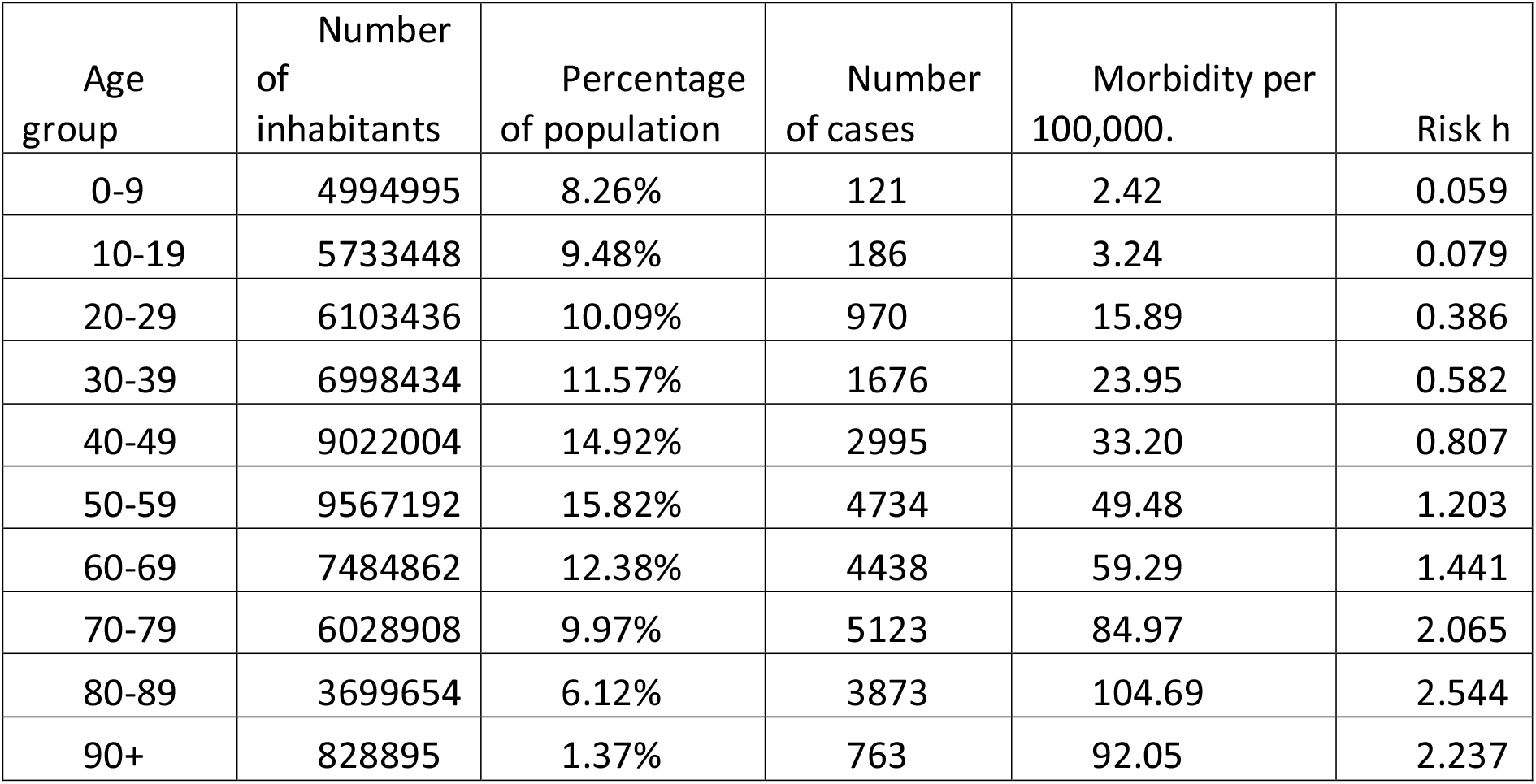
Morbidity and risk by age for the COVID-19 data from Italy.

| Age group | Number of inhabitants | Percentage of population | Number of cases | Morbidity per 100,000. | Risk $h$ |
| --- | --- | --- | --- | --- | --- |
| 0-9 | 4994995 | 8.26% | 121 | 2.42 | 0.059 |
| 10-19 | 5733448 | 9.48% | 186 | 3.24 | 0.079 |
| 20-29 | 6103436 | 10.09% | 970 | 15.89 | 0.386 |
| 30-39 | 6998434 | 11.57% | 1676 | 23.95 | 0.582 |
| 40-49 | 9022004 | 14.92% | 2995 | 33.20 | 0.807 |
| 50-59 | 9567192 | 15.82% | 4734 | 49.48 | 1.203 |
| 60-69 | 7484862 | 12.38% | 4438 | 59.29 | 1.441 |
| 70-79 | 6028908 | 9.97% | 5123 | 84.97 | 2.065 |
| 80-89 | 3699654 | 6.12% | 3873 | 104.69 | 2.544 |
| 90+ | 828895 | 1.37% | 763 | 92.05 | 2.237 |

Next, for the point where the epidemic starts to decline, we can assess the proportions of immune subpopulations by age using the assumptions that 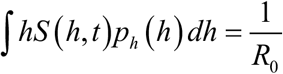 and *F(t*) ≈ 0,5145. These results are presented in Table 2. Notably, the distribution is highly non-uniform, and the average proportion of people with immunity is approximately 43% instead of 60%, as estimated with the homogeneous model.

**Table 2.**
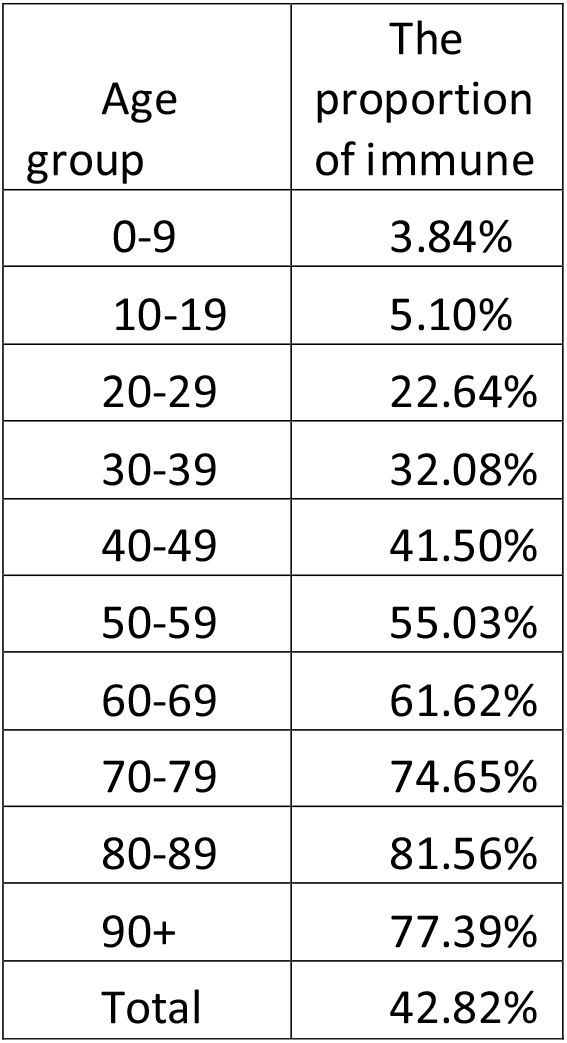
The minimum proportion of infected people by age needed for the epidemic to start declining. The estimate is based on COVID-19 from Italy.

| Age group | The proportion of immune |
| --- | --- |
| 0-9 | 3.84% |
| 10-19 | 5.10% |
| 20-29 | 22.64% |
| 30-39 | 32.08% |
| 40-49 | 41.50% |
| 50-59 | 55.03% |
| 60-69 | 61.62% |
| 70-79 | 74.65% |
| 80-89 | 81.56% |
| 90+ | 77.39% |
| Total | 42.82% |

## Discussion

Here we looked into the issue of assessing the level of collective immunity ^16-19^, which would be safe for lifting anti-epidemic restrictions. We assumed that the basic reproductive number would be approximately 2.5 without restrictions^20-22^. Our assumptions were based on a heterogeneous model which, unlike the commonly used homogeneous models, accounted for the age-related differences in infection risk. We used the COVID-19 data from Italy to generate the estimates.

The heterogeneous model yielded the minimum level of 43% for collective immunity to stop the epidemic, which is different from the 60% level predicted by a homogeneous model. This difference is related to the highly uneven morbidity across the age groups, with the elderly being the most affected. The estimated 43% correspond to all forms of the infectious process development, including both symptomatic and asymptomatic cases. Given the estimated proportion of manifested cases of 50% of the total, we estimate the minimum proportion of Italians who have to suffer coronavirus infection before the nation can return to the normal lifestyle as 20%.

For a better assessment of the epidemic dynamics, additional sources of heterogeneity should be considered ^23,24^, including heterogeneity by social and professional activity, heterogeneity associated with sex and heterogeneity associated with individual resistance to the pathogen. These sources of heterogeneity could lower the minimum collective immunity even further. For example, for Italy this number could be lower than 20%.

## Data Availability

This is a modeling epidemiological study that uses online data repository.

https://www.europeandataportal.eu/en/highlights/covid-19

## References

1. World Health Organization. Coronavirus disease 2019 (COVID-19): situation report, 72. 2020.

2. Prompetchara E, Ketloy C, Palaga T. Immune responses in COVID-19 and potential vaccines: Lessons learned from SARS and MERS epidemic. Asian Pac J Allergy Immunol 2020; 38(1): 1–9.

3. Ahmed SF, Quadeer AA, McKay MR. Preliminary identification of potential vaccine targets for the COVID-19 coronavirus (SARS-CoV-2) based on SARS-CoV immunological studies. Viruses 2020; 12(3): 254.

4. Le TT, Andreadakis Z, Kumar A, et al. The COVID-19 vaccine development landscape. Nat Rev Drug Discov 2020.

5. Lurie N, Saville M, Hatchett R, Halton J. Developing Covid-19 vaccines at pandemic speed. New England Journal of Medicine 2020.

6. Li MY, Smith HL, Wang L. Global dynamics of an SEIR epidemic model with vertical transmission. SIAM Journal on Applied Mathematics 2001; 62(1): 58–69.

7. Zhang J, Ma Z. Global dynamics of an SEIR epidemic model with saturating contact rate. Mathematical Biosciences 2003; 185(1): 15–32.

8. Kwok KO, Lai F, Wei WI, Wong SYS, Tang JW. Herd immunity–estimating the level required to halt the COVID-19 epidemics in affected countries. Journal of Infection 2020.

9. Fine P, Eames K, Heymann DL. “Herd immunity”: a rough guide. Clinical infectious diseases 2011; 52(7): 911–6.

10. John TJ, Samuel R. Herd immunity and herd effect: new insights and definitions. European journal of epidemiology 2000; 16(7): 601–6.

11. Vattay G. Predicting the ultimate outcome of the COVID-19 outbreak in Italy. arXiv preprint arXiv:200307912 2020.

12. de Vlas SJ, Coffeng LE. A phased lift of control: a practical strategy to achieve herd immunity against Covid-19 at the country level. medRxiv 2020.

13. Liu K, Chen Y, Lin R, Han K. Clinical features of COVID-19 in elderly patients: A comparison with young and middle-aged patients. Journal of Infection 2020.

14. Lee P-I, Hu Y-L, Chen P-Y, Huang Y-C, Hsueh P-R. Are children less susceptible to COVID-19? Journal of Microbiology, Immunology, and Infection 2020.

15. Kermack WO, McKendrick AG. A contribution to the mathematical theory of epidemics. Proceedings of the royal society of london Series A, Containing papers of a mathematical and physical character 1927; 115(772): 700–21.

16. Rampini AA. Sequential Lifting of COVID-19 Interventions with Population Heterogeneity: National Bureau of Economic Research, 2020.

17. Favero CA, Ichino A, Rustichini A. Restarting the economy while saving lives under Covid-19. Available at SSRN 3580626 2020.

18. Zhigljavsky A, Whitaker R, Fesenko I, Kremnizer K, Noonan J. Comparison of different exit scenarios from the lock-down for COVID-19 epidemic in the UK and assessing uncertainty of the predictions. arXiv preprint arXiv:200404583 2020.

19. Dolbeault J, Turinici G. Heterogeneous social interactions and the COVID-19 lockdown outcome in a multi-group SEIR model. arXiv preprint arXiv:200500049 2020.

20. Kucharski AJ, Russell TW, Diamond C, et al. Early dynamics of transmission and control of COVID-19: a mathematical modelling study. The lancet infectious diseases 2020.

21. Viceconte G, Petrosillo N. COVID-19 R0: Magic number or conundrum? Infectious disease reports 2020; 12(1).

22. Bar-On YM, Flamholz AI, Phillips R, Milo R. SARS-CoV-2 (COVID-19) by the numbers. arXiv preprint arXiv:200312886 2020.

23. Gomes MGM, Aguas R, Corder RM, et al. Individual variation in susceptibility or exposure to SARS-CoV-2 lowers the herd immunity threshold. medRxiv 2020.

24. Zhigljavsky A, Whitaker R, Fesenko I, et al. Generic probabilistic modelling and non-homogeneity issues for the UK epidemic of COVID-19. arXiv preprint arXiv:200401991 2020.

